# Cell-Free DNA in Blood Reveals Significant Cell, Tissue and Organ Specific injury and Predicts COVID-19 Severity

**DOI:** 10.1101/2020.07.27.20163188

**Authors:** Alexandre Pellan Cheng, Matthew Pellan Cheng, Wei Gu, Joan Sesing Lenz, Elaine Hsu, Erwin Schurr, Guillaume Bourque, Mathieu Bourgey, Jerome Ritz, Francisco Marty, Charles Y. Chiu, Donald Cuong Vinh, Iwijn De Vlaminck

## Abstract

COVID-19 primarily affects the lungs, but evidence of systemic disease with multi-organ involvement is emerging. Here, we developed a blood test to broadly quantify cell, tissue, and organ specific injury due to COVID-19, using genome-wide methylation profiling of circulating cell-free DNA in plasma. We assessed the utility of this test to identify subjects with severe disease in two independent, longitudinal cohorts of hospitalized patients. Cell-free DNA profiling was performed on 104 plasma samples from 33 COVID-19 patients and compared to samples from patients with other viral infections and healthy controls. We found evidence of injury to the lung and liver and involvement of red blood cell progenitors associated with severe COVID-19. The concentration of cfDNA correlated with the WHO ordinal scale for disease progression and was significantly increased in patients requiring intubation. This study points to the utility of cell-free DNA as an analyte to monitor and study COVID-19.

## INTRODUCTION

The Coronavirus Disease-19 (COVID-19) pandemic is a major global health crisis. COVID-19 is a complex disease with diverse clinical features, ranging from asymptomatic infection to acute respiratory distress syndrome (ARDS) and multi-organ dysfunction. There is an urgent need for predictive biomarkers of COVID-19 severity detectable early in disease onset, and improved understanding of the pathogenesis of COVID-19. Here, we have investigated the utility of circulating cell-free DNA (cfDNA) in blood as an analyte ***i)*** to broadly monitor cell, tissue, and organ injury due to COVID-19, ***ii)*** to assess disease severity and predict disease outcomes, and ***iii)*** to elucidate the multi-organ involvement that characterizes COVID-19.

Autopsy studies indicate a broad organotropism for the SARS-CoV-2 virus beyond the lungs [1], [2]. Detection of the virus in the kidneys, heart, liver, brain and blood of many patients has been reported [2], [3]. The significant viral burden in the kidney seen in some patients may help explain the increased risk of acute kidney injury in patients with COVID-19. Damage to endothelial cells may contribute to COVID-19 coagulopathy and prothrombic state [4]–[8].

Initial reports have primarily described COVID-19 as a disease affecting tissues expressing ACE-2 [9]. However, there are emerging data that SARS-CoV-2 infection may also be accompanied by hematological derangements [10]–[13]. In addition, a dysregulated immune response to SARS-CoV-2 can occur, contributing to the development of ARDS, systemic tissue injury, and multi-organ failure [14]. A strong association between increased cytokine profiles and the severe deterioration of some patients has been observed [15]. In children, a multisystem inflammatory syndrome linked to recent SARS-CoV-2 infection is reported [16]. Given the disparate clinical manifestations and potential complications of COVID-19, there is an urgent need for tests that can quantify injury to multiple tissues simultaneously to monitor patients, analyze disease pathogenesis, predict clinical outcomes, and guide clinical management in patients with COVID-19.

Since the advent of cfDNA based noninvasive prenatal testing, myriad applications of cfDNA in diagnostic medicine have been established [17]–[19]. These short fragments of circulating DNA are the debris of dead cells from across the body. The value of cfDNA as a quantitative marker of tissue and organ injury was first recognized in solid-organ transplantation, where the level of transplant donor derived cfDNA in the blood is now widely used as a marker of transplant rejection [20]–[22]. More recently, several approaches have been developed to quantify the tissues-of-origin of cfDNA and thus monitor injury to any cell, tissue or organ type [23]–[27]. This is achieved by profiling epigenetic marks within cfDNA by quantitative molecular measurement technologies such as DNA sequencing. Here, we tested the hypothesis that cfDNA tissues-of-origin profiling enables the identification of specific tissue or cell types that are directly or indirectly targeted and injured throughout COVID-19 pathogenesis. We studied two independent patient cohorts, and found evidence of significant injury to the liver, lung, and kidney associated with COVID-19. We further observed a striking increase, both in terms of proportion and total abundance, of cfDNA derived from red blood cell precursors when compared to patients infected with other RNA viruses and healthy controls. Last, the total burden of cfDNA correlated with the WHO ordinal scale for disease progression, with an increase in cfDNA being strongly associated with admission to the intensive care unit and need for mechanical ventilation. Thus, cfDNA can provide a marker of disease severity as well as a prognostic tool that is straightforward to adopt.

## RESULTS

We tested the utility of cfDNA to quantify cell, tissue, and organ specific injury associated with COVID-19 in two independent patient cohorts from two different hospitals in North America (**Fig. 1A, supplementary table 1**,**2**). We assayed a total of 104 plasma samples from 33 patients across these cohorts. We performed shotgun DNA sequencing after bisulfite treatment to determine the tissues-of-origin of cfDNA isolated from all plasma samples by methylation profiling. We obtained 62 ± 35 million (mean ± standard deviation) paired-end reads per sample, leading to a per-base genome coverage of 1.3 ± 0.8. We verified that we achieved a high bisulfite conversion efficiency for all samples (0.996 ± 0.005, Methods). To determine the cell, tissue, and organ types that contribute cfDNA to the mixture in blood (Methods), we analyzed plasma cfDNA methylation profiles against a reference set of 147 cell, tissue, and organ types using previously described bioinformatic approaches (**Fig. 1B**,**C, supplementary data 1**, Methods) [25].

**Figure 1.**
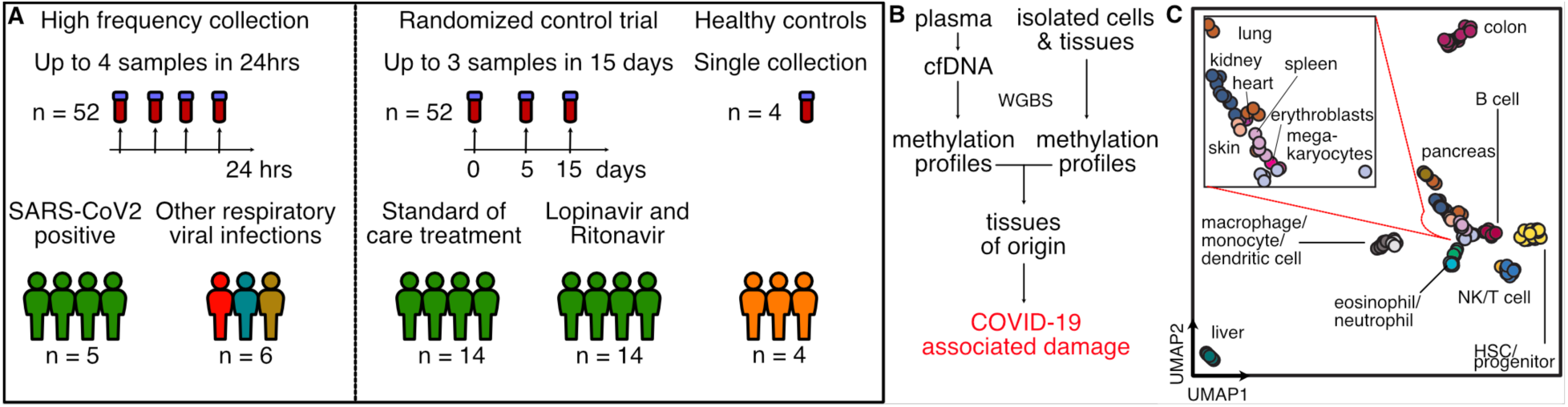
Study design. **A)** Two independent cohorts were used in our study: First, a high frequency collection cohort with 5 SARS-CoV-2 patients (n = 52 samples) and 6 SARS-CoV-2 negative, RNA-virus positive patients (n = 6 samples). Second, a randomized control trial of 28 SARS-CoV-2 patients with plasma at serial time points (n = 52 samples). 4 healthy individuals volunteered plasma for cell-free DNA analysis. **B)** Experimental workflow. cfDNA is extracted from plasma and whole-genome bisulfite sequencing is performed. In parallel, methylation profiles of cell and tissue genomes are obtained from publicly-available databases. cfDNA methylation profiles are compared to those of cell and tissue references to infer relative contributions of tissues to the cfDNA mixtures. **C)** UMAP of differentially methylated regions for isolated cell and tissue types used as a reference.

### Temporal dynamics of cell-free DNA tissues-of-origin in plasma of COVID-19 patients

We first assayed 52 serial samples collected at short time intervals from five adult patients with COVID-19 that were treated at University of California, San Francisco (UCSF) Medical Center (median of 8 samples per patient [range 6-18]). These plasma samples were residual from clinical testing and were collected from this group of patients over a treatment time-period of up to 14 days with up to four samples collected within 24 hours (median time between consecutive collections of 13 hours [range 5-64]). These samples allowed us to study dynamic changes in cfDNA profiles in patients diagnosed with and treated for COVID-19 (**Fig. 2A**). Treatments included standard of care (n=2), remdesivir (n=1), hydroxychloroquine (n=1), or a combination of remdesivir, hydroxychloroquine, azithromycin and tocilizumab (n=1). In addition to plasma from COVID-19 patients, we performed cfDNA tissues-of-origin profiling for six samples collected from patients with other respiratory viral infection treated at the same hospital, including influenza B (n=2), Metapneumovirus (n=1), Coronavirus HKU1 (n=1), Coronavirus NL63 (n=1) and Respiratory syncytial virus B (n=1) (**Fig. 2B**).

**Figure 2.**
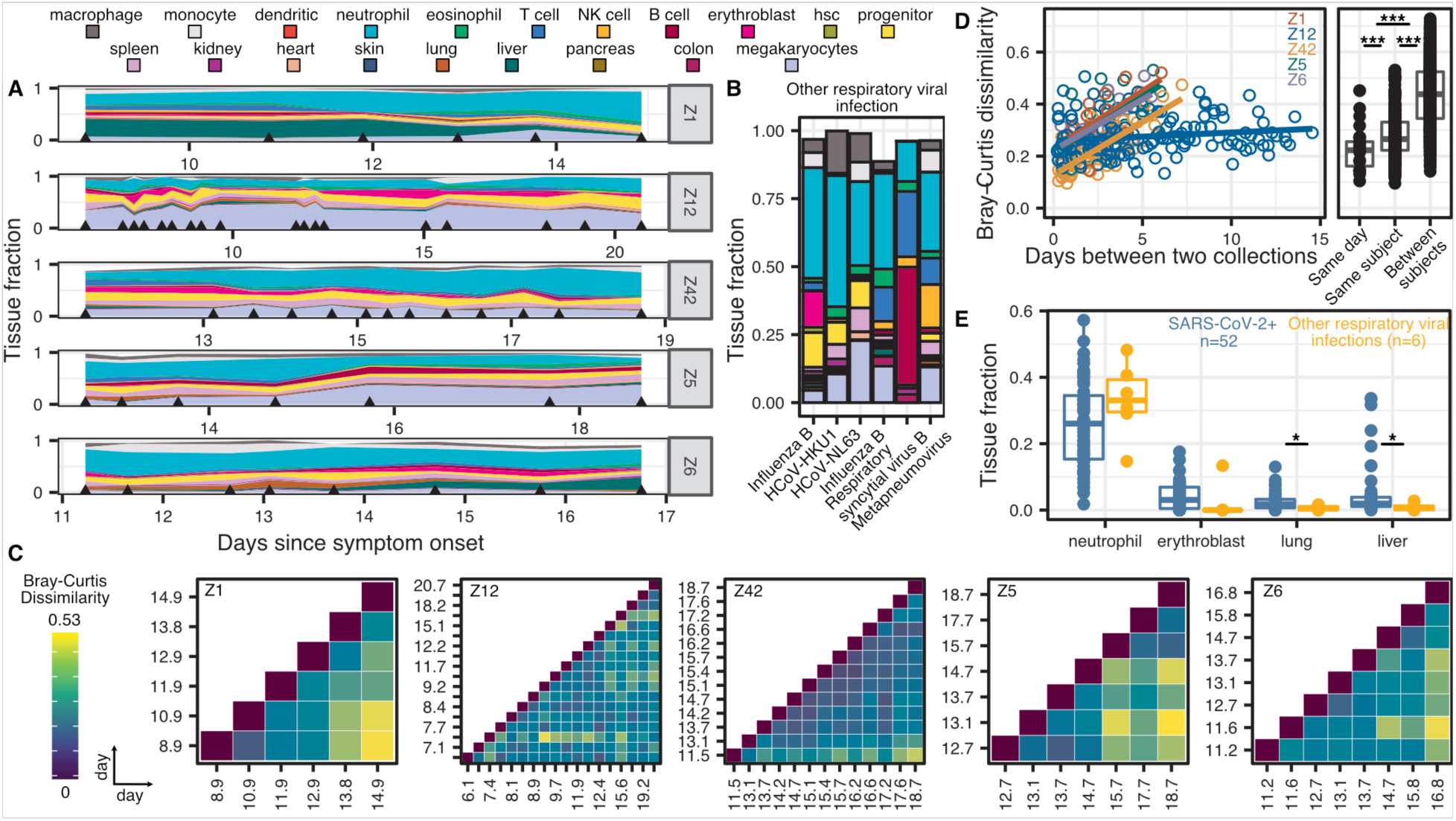
High frequency sample collection cohort at UCSF. **A-B)** Patient-specific relative tissue contributions for SARS-CoV-2 patients (**A**) and other RNA-virus infection patients (**B**). Triangles indicate sampling times. **C)** Heatmaps of Bray-Curtis dissimilarity. **D)** Scatterplot of patient-specific Bray-Curtis dissimilarity (left) and boxplot of Bray-Curtis dissimilarity between cfDNA tissue proportions from samples collected from either the same day (within 24 hours), the same person (but not within 24 hours), or from all patients (right). **E)** Comparison of tissue fraction of four cell and tissue types (neutrophil, erythroblast, lung and liver) between SARS-CoV-2 positive patients and other RNA-virus positive patients. * : p-value < 0.05; ** : p-value < 0.01; *** : p-value < 0.001 (p-values calculated using a Wilcoxon test)

We plotted the relative abundance of cfDNA derived from different cell, tissue, and organ types and found that differences in cfDNA profiles between individuals were larger than differences within individuals over the sampling period. For subjects Z1, Z5, Z6, and Z42 but not Z12, we observed gradual changes in the tissues-of-origin profiles over sampling periods of six to seven days. We used the Bray Curtis dissimilarity to quantify the inter and intra-individual differences in cfDNA profiles (**Fig. 2C-D)**. This analysis confirmed the visual appearance of the tissues-of-origin profiles in Figure 2A and demonstrated that the largest differences in cfDNA were found for samples collected from different individuals. Within subjects, smaller differences were observed for samples collected on the same day **(Fig. 2D**). Last, the Bray Curtis dissimilarity increased with time interval between samples for patients Z1, Z5, Z6, and Z42 but not for Z12. Together these analyses indicate that cfDNA profiles are subject specific, and that changes in cfDNA tissues-or- origin profiles occur gradually over days and not hours, therefore adequate longitudinal data can be collected every few days.

We next compared the cfDNA tissues-of-origin profiles associated with COVID-19 versus those associated with respiratory infection with other viruses (**Fig. 2E, supplementary table 3**). We found significant increases in the relative proportion of lung specific cfDNA in the blood of COVID-19 patients, which was likely related to COVID-19 associated tissue injury (2.5% vs 0.6%, p-value = 0.019, Wilcoxon). We found a similar association with liver-derived cfDNA (5.0% vs 0.9%, p-value = 0.025, Wilcoxon), and this was validated by the elevated liver function tests in 4 of 5 COVID-19 patients. Strikingly, we also observed an increase in the relative proportion of cfDNA derived from erythroblasts in the blood of COVID-19 patients compared to the control group (75% vs 17% samples with an erythroblast fraction greater than 0, p-value = 0.003, 2-sample proportions test, **Fig. 2E, supplementary figure 1**). Erythroblasts are nucleated cells typically in the adult bone marrow from which red blood cells develop. The increase in cfDNA derived from red blood progenitor cells seen here may be an indirect consequence of the hypoxemia and/or cytokine-mediated anemia that characterize severe COVID-19, or may indicate a more direct involvement of coronavirus with red blood cell precursors. We note that erythroblast cfDNA was elevated in a single patient in the control group, who was being treated for recurrent stage IV diffuse large B-cell lymphoma (**Fig. 2 B**,**E**).

### Randomized clinical trial cohort

To test the robustness of these initial observations, we assayed an additional 52 samples collected from 28 patients that were recruited into a randomized control trial at the McGill University Health Centre in Montreal, Canada. Patients were assigned to either an experimental antiviral therapy consisting of a combination of Lopinavir and Ritonavir (brand name Kaletra) or to the standard of care. Of these patients, 14 were treated with the Lopinavir/Ritonavir, and 14 were treated with the standard of care. Of the 28 patients, 21 were discharged after treatment, one patient remains hospitalized as of July 19^th^, 2020, and six patients died. Serial samples were collected from these patients at three predetermined time points: days 1, 5, and 15 after enrollment in the clinical trial, provided they remained hospitalized on the days of collection (**Fig. 3A**). We determined the relative abundance of tissue-specific cfDNA using the approaches described above. In addition, we quantified the absolute concentration of tissue-specific cfDNA by multiplying the proportion of tissue-specific cfDNA with the concentration of total cfDNA (Methods).

**Figure 3.**
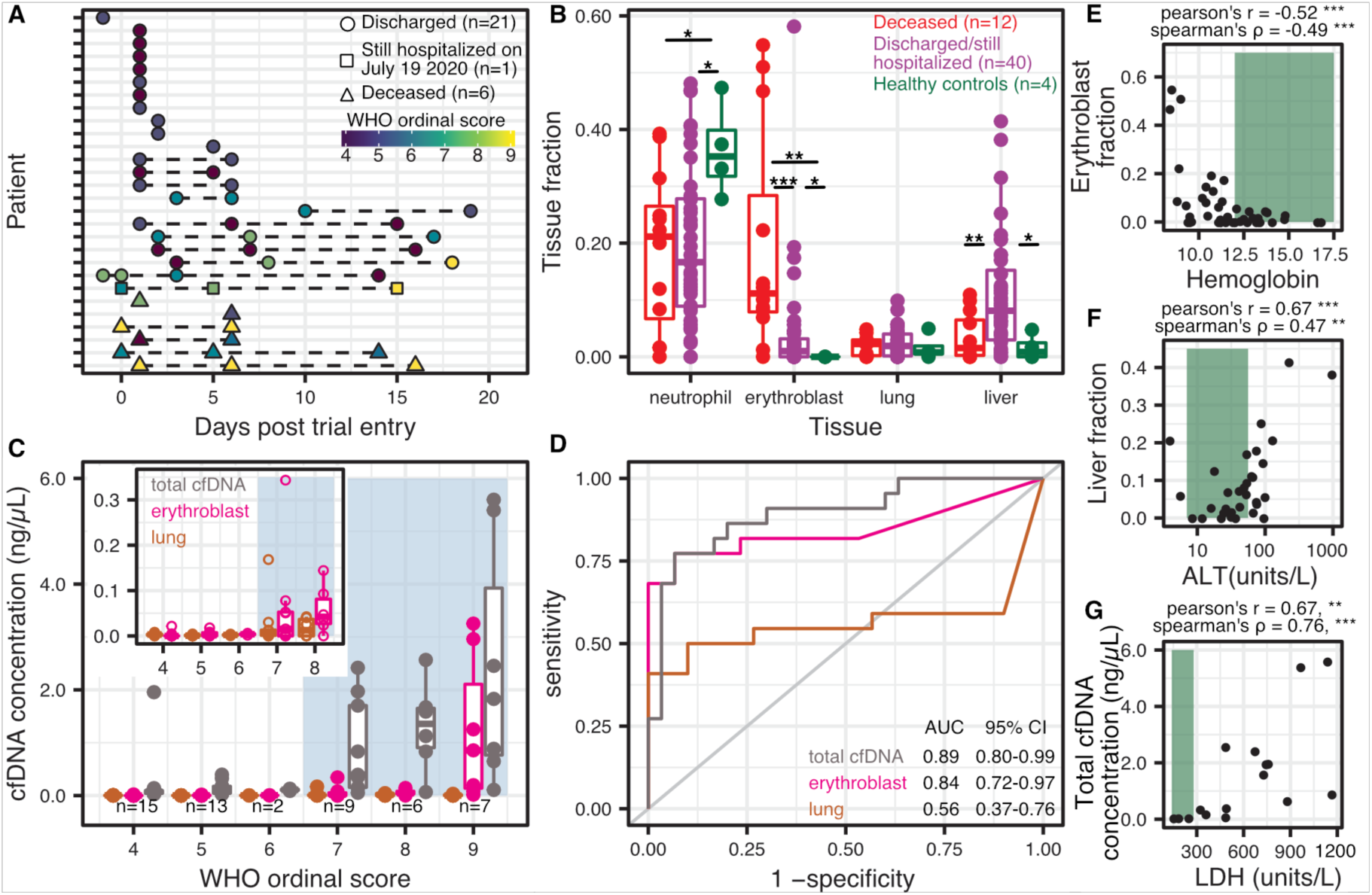
Randomized control trial cohort from MUHC. **A)** Patient sample-collection map by day of enrollment into the study. **B)** Relative proportion of cfDNA derived from four cell and tissue types (neutrophil, erythroblast, lung, liver) by hospitalization status (p-values calculated using a Wilcoxon test). **C)** Absolute cfDNA concentrations compared to the WHO ordinal scale for COVID progression. Blue shading indicates ordinal scores requiring admittance to the intensive care unit (ICU) **D)** Receiver operating characteristic analysis of the performance of absolute cfDNA concentration of different tissues (lung, erythroblast and total) in distinguishing patients presenting with ordinal scales from 4-6 (hospitalized) and 7-9 (hospitalized in the ICU). **E-G** Scatterplot comparisons between relative proportions of erythroblast cfDNA fraction and hemoglobin (**E**), liver cfDNA fraction and alanine aminotransferase (ALT) (**F**) and total cfDNA concentration and lactase dehydrogenase (LDH) (**H**). Green shading indicates normal levels. * : p-value < 0.05; ** : p-value < 0.01; *** : p-value < 0.001.

We first compared the cfDNA tissues-of-origin profiles measured for these patients with the tissues-of-origin profiles for four healthy subjects (**Fig. 3B, supplementary figure 2**). We found that 62% of samples from patients with COVID-19 had a higher concentration of lung cfDNA than the highest concentration measured for a healthy individual (p-value = 0.017, 2-sample proportions test). In addition, hospitalized patients with COVID-19 had both an elevated relative and absolute burden of cfDNA derived from the liver (liver fraction 9.1 vs 1.6%, p-value = 0.054, and 0.051 ng/µL vs 0.00029 ng/µL, p-value = 0.010, Wilcoxon). In addition to these tissue-specific features, we again observed a significant increase in cfDNA derived from erythroblast cells for COVID-19 patients compared to healthy controls (7.7% vs 0%, p-value = 0.027, Wilcoxon; 65% vs 0% of samples showing erythroblast fraction greater than 0, p-value = 0.0099, 2-sample proportions test, **Fig. 3B**). We evaluated the temporal dynamics of the contribution of different cell and tissue types to the mixture in plasma of COVID-19 patients and observed a slow recovery in tissue injury and a slow increase in the contribution of cfDNA derived from erythroblasts (**supplementary figure 3**).

We then compared cfDNA signatures for COVID-19 patients as function of disease severity, and found that erythroblast cfDNA proportions at any timepoint are predictive of in-hospital mortality (19.6% vs 4.1%, p-value = 0.0004, Wilcoxon). Receiver operating characteristic (ROC) analysis of the performance of the relative proportion of Erythroblast derived DNA to predict COVID-19 mortality yielded an area under the curve (AUC) of 0.83 (95% CI 0.69-0.98, [deceased n = 12; hospitalized or discharged n = 40]). Additionally, our analysis revealed that kidney cfDNA was significantly elevated in COVID-19 patients who eventually died (1.8% vs 0.5% vs 0.005% between deceased, non-deceased and healthy controls, p-value = 0.0018 between deceased and non-deceased COVID-19 patients).

We then compared the cfDNA tissues-of-origin profiles to the WHO clinical progression scale for COVID-19 [28] (**Fig. 3C**). We found a strong association between the total cfDNA concentrations isolated from plasma and the WHO clinical progression scores (**Fig. 3C**,**D**). Notably, a clinical score of 7 or greater (indicating the need for admission to the intensive care unit and invasive mechanical ventilation), was associated with a sharp increase in the total burden of cfDNA (**Fig. 3C**,**D**, mean 1.5 ng/µL vs 0.16 ng/µL, between clinical scores from 7 to 9 and 4 to 6, respectively; p-value = 1.5−10^−6^, Wilcoxon). ROC analysis of cfDNA concentrations to predict ordinal scores revealed AUCs of 0.89 (95% CI 0.80-0.99), 0.84 (95% CI 0.72-0.97) and 0.56 (95% CI 0.37-0.76) for total, erythroblast and lung cfDNA, respectively. Furthermore, samples taken from patients with a clinical score of 9 (use of extracorporeal membrane oxygenation [ECMO]) had significantly higher erythroblast-derived cfDNA than patients with a clinical score of 7-8 (1.23 ng/µL vs 0.06 ng/µL, p-value = 0.006, Wilcoxon). Patients on ECMO tend to bleed and require additional blood volumes, which may contribute to the increased erythroblast signal. However, erythroblast-derived cfDNA was significantly increased in patients with a clinical score of 7 or higher as well (**Fig. 3C**,**D**, mean 0.43 ng/µL vs 0.003 ng/µL, p-value = 1.83−10^−5^, Wilcoxon).

Erythroblast and liver cfDNA contributions correlated with clinical metrics for anemia and liver damage, respectively (**Fig3. E-G**). We observed significant negative correlations between the proportion of erythroblast cfDNA and hematocrit and hemoglobin (Pearson’s R (R) = −0.51, Spearman’s ρ (ρ) = −0.37 and R = −0.52, ρ = −0.49, respectively). Similarly, we found positive correlations between the proportion of liver-derived cfDNA and alanine aminotransferase (ALT) and aspartate transaminase (R = 0.63, ρ = 0.47 and R = 0.76, ρ = 0.24, respectively). We did not observe a correlation between kidney-derived cfDNA and serum creatinine (R = 0.05, ρ = 0.09). We found similar results when comparing the tissue-derived cfDNA concentration to these clinical markers (erythroblast cfDNA concentration vs hematocrit and hemoglobin: R = −0.42, ρ = −0.32 and R = −0.38, ρ = −0.45, respectively. Liver cfDNA concentration vs ALT and AST: R = 0.84, ρ = 0.52 and R = 0.20, ρ = 0.23, respectively. Kidney cfDNA concentration vs creatinine: R = 0.56, ρ = 0.20).

Recent papers from Yan *et al*. and Zhou *et al*. identified lactate dehydrogenase (LDH) as a strong predictor of COVID-19 outcome [29], [30]. LDH is found in virtually all cells and is a commonly used biomarker for tissue damage and hemolysis [31]–[33]. We found significant correlation between LDH and the proportion of erythroblast-derived cfDNA (R = 0.64, ρ = 0.65), and between LDH and total cfDNA (R = 0.67, ρ = 0.76). Together, these data suggest that cfDNA tissues-of-origin can be applied to resolve the specific tissues contributing to non-specific detection of LDH in blood.

Finally, we found no differences between lung, liver, kidney or erythroblast-derived cfDNA for patients receiving standard of care, or the experimental lopinavir/ritonavir treatment (**supplementary figure 4**). These data are in line with the results of recent clinical trials that treatment with lopinavir/ritonavir is not significantly different from standard of care treatment for COVID-19 [34], [35].

## DISCUSSION

We find significant support for the utility of cfDNA profiling as a prognostic tool for the early detection and monitoring of cell and tissue injury associated with COVID-19. A minimally invasive molecular blood test that can inform cell, tissue and organ specific injury due to COVID-19 has the potential to alleviate the impact of the COVID crisis ***i*)** by providing quantifiable prognostic parameters and a more granular assessment of clinical severity at the time of presentation; and ***ii)*** by providing a surrogate biomarker that can be included in clinical trials of candidate COVID-19 treatments.

In line with the diverse clinical manifestations of COVID-19, we find evidence for lung, liver and kidney injury in hospitalized patients with COVID-19. While lung-derived cfDNA was elevated in COVID-19 patients, we did not find it to be a major contributor to plasma cfDNA. The level of lung specific cfDNA in plasma was similar to the levels observed in lung transplant patients that suffer acute lung transplant rejection [20] and lung cancer patients [36], [37]. We observed a striking correlation between the total abundance of circulating cfDNA in plasma and the WHO ordinal scale for disease progression. We propose that the total abundance of cfDNA, which can be measured within one hour at a low cost, can be used in the context of clinical trials and patient management in the near term.

In addition to the practical application of cfDNA profiling to patient monitoring and COVID-19 risk stratification, the cfDNA methylation assay and data reported may help elucidate aspects of COVID-19 pathogenesis. The most significant cfDNA signature observed in the two cohorts relative to controls was an increase in cfDNA derived from erythroid or red blood progenitor cells. Given that cfDNA is estimated to have a half-life of about 1 hour [38] and that the proportion of the erythroid lineage was relatively stable over several days, the elevated erythroid cfDNA is likely due to a continuous increased erythroid turnover. In support of elevated erythroid turnover and production, two recent studies have identified red blood cell distribution width (RDW), a measure of the variation in size of red blood cells (RBCs), as an important prognostic predictor for severe COVID-19 [15], [16]. The increased RDW was speculated to be associated with increased turnover of RBCs since increased reticulocytes or newly formed RBCs have a wider diameter [16]. However, our analysis demonstrated that there was no association with RDW and patient outcomes (mean 15.4 vs 14.0 between deceased and discharged or hospitalized, p-value = 0.2, Wilcoxon) and that erythroblast cfDNA was not strongly correlated with RDW (R = 0.26, ρ = 0.13 [with data from UCSF and MUHC]).

Increased erythroid turnover may be due to erythroid destruction as the primary driver, followed by compensatory production, and is supported by anemia (Hgb <13.5 g/dL for men and Hgb < 12 for women) found in 26 of 33 COVID-19 patients across both studies. Possible mechanisms include: ***i)*** excessive inflammation and cytokine storm [39], [40], ***ii)*** hemophagocytosis in relation to inflammation [41], and ***iii)*** consumption in microthrombi [6]–[8], [10]. We note that 18 of 33 patients in all studies, C-reactive protein (CRP) was elevated (> 10 mg/L). It is notable however that megakaryocytes proportions were not increased in either cohort and would not support microthrombi as the predominant reason for increased erythroid turnover. Alternatively, past work has shown that angiotensin II regulates normal erythropoiesis and stimulates early erythroid proliferation through unclear downstream mechanisms [42]–[44]. The binding of SARS-CoV-2 to the host ACE2 may dysregulate erythropoiesis through the downstream angiotensin II pathway. The significant increase in cfDNA derived from red blood progenitor cells, may alternatively be due to injury to red cell precursors [45], through direct or indirect processes. These hypotheses are further testable through various routes, including comprehensive evaluation of erythrocytosis in patients with COVID-19, for example through evaluation of circulating reticulocytes and evaluation of the bone marrow; these measures were not systematically in place during the initial rapid wave of the pandemic and were not implemented in this study.

This study has several limitations. First, we assayed samples from only hospitalized patients, and we have not evaluated cfDNA profiles for mild COVID-19 cases. Second, while this study spans two independent cohorts, with patient groups that are genetically and geographically unrelated, the overall sample size and patient numbers may not be sufficient to generalize our findings to the entire spectrum of COVID-19 cases. Nonetheless, our analysis of cfDNA tissues of origin can provide immediate insights into the dynamics and pathogenesis of COVID-19. Last, the resolution of our measurements is limited by the availability of isolated cells and tissue methylation patterns. Our current reference dataset does not include all known human cell types and tissue types. Therefore, we are not sensitive to those rarer tissues that may play a role in the pathogenesis of COVID-19. More comprehensive investigations are therefore needed to confirm and further refine the observations reported here.

In summary, we report the application of cfDNA profiling to quantify cellular and tissue specific injury due to COVID-19.

## MATERIALS AND METHODS

### High frequency sampling

Clinical samples from UCSF were processed through protocols approved by the UCSF Institutional Review Board (protocol number 10-00476, 18-25287). Residual plasma was collected as part of routine clinical testing and stored at 4 °C for up to 5 days and subsequently stored at -80 °C until batched extraction. Plasma was initially isolated from blood by the clinical laboratory after centrifugation at approximately 800g for 10 minutes. After storage, the plasma was centrifuged at 16,000g for 10 minutes. cfDNA extraction was performed according to manufacturer recommendations (Qiagen MinElute Circulating Nucleic Acid Kit, reference #55204 or Qiagen EZ1 Virus Mini Kit v2.0 955134) at 0.4-1 mL plasma input.

### Randomized clinical trial

Individuals diagnosed with COVID-19 were recruited to a randomized, controlled clinical trial at the McGill University Health Center, where they received either Lopinavir/ritonavir, or standard-of-care (https://clinicaltrials.gov/ct2/show/NCT04330690). Blood samples were collected under MUHC Research Ethics Board protocol 10-256 through standard venipuncture in standard blood collection tubes and immediately centrifuged at 850g for 10 minutes. The supernatant is then transferred to new tubes, and centrifuged at 16,000g for 10 minutes. Plasma-containing supernatant is collected and stored in DNA cryostorage vials (Eppendorf, reference #0030079400) at -80 °C. Plasma was shipped overnight on dry ice from the McGill University Health Center (Montreal, Canada) to Cornell University (Ithaca, United-States). Plasma was stored at -80 °C until used for cfDNA extraction. cfDNA extraction was performed according to manufacturer recommendations (Qiagen Circulating Nucleic Acid Kit, reference #55114).

### Healthy controls

Volunteers were recruited for blood donations through a protocol approved by the Cornell Institutional Review Board (protocol number 1910009101). Blood was collected in K2 EDTA tubes (BD, reference #366643) and immediately centrifuged at 1600g for 10 minutes. The supernatant was transferred to new tubes, and centrifuged at 16,000g for 10 minutes. Supernatant is then stored in DNA cryostorage vials (Thermo Scientific #363401) at -80 °C until cfDNA extraction. cfDNA extraction was performed according to manufacturer recommendations (Qiagen Circulating Nucleic Acid Kit, reference #55114).

### Whole genome bisulfite sequencing

Bisulfite treatment of DNA converts cytosine residues to uracil but leaves methylated cytosines unaffected [46]. DNA sequencing of bisulfite-treated cfDNA can be used to reveal methylation patterns with single nucleotide resolution. Because these patterns are cell, tissue, and organ types specific, they can inform the origins of cfDNA. Following treatment with bisulfite, whole-genome sequencing (WGS) libraries were prepared according to manufacturer’s protocols (Zymo EZ Methylation-Gold kit, #D5005 and Swift Biosciences Accel-NGS Methyl-Seq DNA Library Kit #30024) using a dual indexing barcode strategy (Swift biosciences #38096, NEBNext Multiplex Oligos for Illumina E7500L, or custom primers). Paired-end DNA sequencing was performed on the Illumina NextSeq 500 (2×75bp) at Cornell University or the Illumina NovaSeq (2×150bp) at University of California San Francisco. Resulting paired-end fastq files were trimmed to 75bp for downstream analysis.

### Human genome alignment

Adapter sequences were trimmed using BBDUK (BBTools software suite [47]). Resulting sequences were aligned to the human genome (version hg19) and deduplicated using Bismark [48]. Alignment files were filtered with a minimum mapping quality of 10 using SAMtools [49].

### Reference methylomes and tissues of origin

Reference methylation profiles were obtained from publicly available datasets and international epigenetic consortium projects (**supplementary data 1**) and processed as previously described [25]. Briefly, files were downloaded and normalized to a standard 4 column BED format at single nucleotide resolution using hg19 coordinates. Differentially methylated regions (DMRs) were found using Metilene [50]. Methylation densities within these DMRs were averaged. Tissues with methylation profiles highly dissimilar from the same tissues were removed. cfDNA methylation densities were extracted using Bismark [48] and averaged over the DMRs. Tissues of origin were deconvoluted using a non-negative least squares approach.

### cfDNA concentration measurement - MUHC patients

Plasma samples were processed in batches of 4 to 10 alongside a control containing 8 µL of approximately 150 ng/µL of synthetic oligos. DNA concentration measurements were performed after cfDNA extraction (Qubit Fluorometer 3.0) and the normalized concentration was calculated by multiplying the sample’s concentration by the input/output ratio of the control.

### Depth of coverage

The depth of DNA sequencing coverage was calculated by dividing the number of mapped nucleotides to the autosomal chromosomes to the size of the non-N hg19 autosomal genome.

### Bisulfite conversion efficiency

The bisulfite conversion efficiency achieved in experiments was estimated using MethPipe [51] by calculating the reported methylation density of cytosines present at C[A/T/G] dinucleotides, which are rarely methylated in mammalian genomes.

### Quality control filtering

Samples from the high frequency sampling cohort were selected for analysis if 10 or more spike-in molecules were identified after sequencing and were also filtered for sufficient depth of sequencing (>0.2x human genome). Samples from the randomized control trial cohort were sequenced to a minimum depth of 0.7x human genome coverage. All samples had a minimum bisulfite conversion efficiency of 96%.

### Statistical analysis

All statistical analyses were performed in R, version 3.5.0. Groups were compared using the two-sided, nonparametric Wilcoxon test. If the data distributions were zero-skewed, a two-sided, 2-sample proportions test without continuity correction was performed. Boxplots span from the 25th and 75th percentiles. The band in the box indicates the median, lower and higher whiskers extend to the smallest and largest values at most 1.5× IQR of the hinge, respectively.

## Data availability

Genomic data will be hosted on the Sequence Read Archive. The code used to generate figures and analyze primary data is available at www.github.com/alexpcheng/cfDNAme.

## AUTHOR CONTRIBUTIONS

A.P.C., M.P.C., W.G., C.Y.C., D.C.V. and I.D.V. designed the study. M.P.C., W.G., C.Y.C., D.C.V. consented patients and obtained clinical data. J.S.L., E.H, W.G. performed experiments. A.P.C., M.P.C., W.G. and I.D.V. analyzed data. A.P.C., W.G., M.P.C., D.V. and I.D.V. wrote manuscript. All authors provided edits and comments.

## ACKNOWLEDGEMENTS

We thank Dr. Peter Schweitzer and colleagues at the Cornell Genomics Center for help with sequencing assays. We thank Dr. Lynn Johnson and the Cornell Statistical Consulting Unit for help with statistical analysis. We thank Dr. Lucie Roussel, Dr. Marianna Orlova and Pauline Cassart for technical assistance. This work was supported by NIH Grant 1DP2AI138242 (to I.D.V.), R01AI146165 (to I.D.V. and M.P.C), 1R01AI151059 (to I.D.V.), K08-CA230156 (to W.G), R33-AI129455 to C.Y.C, a Synergy award from the Rainin Foundation (to I.D.V.), a SARS-CoV-2 seed grant at Cornell (to I.D.V.), a National Sciences and Engineering Research Council of Canada fellowship PGS-D3 (to A.P.C.), and Burroughs-Wellcome CAMS Award (to W.G.). D.C.V. is supported by a Fonds de la recherche en sante du Quebec Clinical Research Scholar Junior 2 award. C.Y.C. is supported by the California Initiative to Advance Precision Medicine, and the Charles and Helen Schwab Foundation (C.Y.C.).

